# External validation and recalibration of office- and laboratory-based cardiovascular risk scores for prediction of 10-year risk of fatal cardiovascular disease in 112,262 adults in Mexico City

**DOI:** 10.1101/2025.09.07.25335293

**Authors:** Jerónimo Perezalonso-Espinosa, Daniel Ramírez-García, Juan Pablo Díaz-Sánchez, Karime Berenice Carrillo-Herrera, Leslie Alitzel Cabrera-Quintana, Gael Dávila-López, Carlos A. Fermín-Martínez, Andrea Malagón Liceaga, Martín Roberto Basile-Álvarez, Jaime Berumen-Campos, Pablo Kuri-Morales, Roberto Tapia-Conyer, Jesus Alegre-Díaz, Jacqueline A. Seiglie, Neftali Eduardo Antonio-Villa, Omar Yaxmehen Bello-Chavolla

## Abstract

**BACKGROUND:** Cardiovascular disease (CVD) is the leading cause of mortality in Latin America, yet most CVD risk prediction models were developed in high-income countries with limited validations in these settings.

**OBJECTIVES:** To externally validate CVD risk prediction models for 10-year fatal CVD, and recalibrate the Globorisk-fatal model in Mexican population.

**METHODS:** We analyzed 112,262 adults ≥40 years from the Mexico City Prospective Study. Outcomes were restricted to fatal CVD, including myocardial infarction (MI) and stroke, censored at 10 years. CVD risk was estimated using the laboratory- and office-based Framingham, Globorisk, Globorisk-LAC, WHO, and SCORE2 equations. Discrimination was assessed with Harrell’s c-statistic and AUROCs, calibration with mean estimates, slopes, and calibration curves, and overall performance with Brier scores. Sex-specific recalibration of the Globorisk-fatal model was performed using observed 10-year risks.

**RESULTS:** During 10 years of follow-up, 2,429 fatal CVD events were recorded (1,667 MI, 762 stroke). All models showed good discrimination, with c-statistics ranging from 0.761-0.805 in men and 0.797-0.831 in women. The Globorisk-fatal model had the highest c-statistic in women (0.831, 95%CI 0.821-0.841), and the laboratory-based WHO-MI model in men (0.805, 95%CI 0.783-0.827). Despite this, all equations consistently overestimated CVD risk, particularly in women. Calibration analyses revealed systematic overprediction at higher risk levels, more pronounced in men. Recalibration of the Globorisk-fatal model improved agreement between predicted and observed risks, reducing overestimation.

**CONCLUSIONS:** CVD risk models showed good discrimination but consistently overestimated risk in this Mexican cohort. The recalibrated Globorisk-fatal model improves risk estimation of fatal CVD in Mexican adults.

## INTRODUCTION

Cardiovascular diseases (CVDs) are the leading cause of mortality and disability among adults worldwide^1,2^, and low- and middle-income countries carry a disproportionate share of associated premature deaths and economic costs^3^. Although CVD mortality rates have declined in Latin America, population growth and ageing have resulted in an increase of the total number of deaths in the region, and CVDs still accounted for 1.1 million deaths in 2021, underscoring the need for sustained prevention and control strategies^4–6^. A key target of the Sustainable Development Goals is to reduce by one third premature mortality from non-communicable diseases, and to achieve the coverage of least 50% of eligible individuals with drug therapy and counseling to prevent myocardial infarction and stroke^3,7^. Treatment eligibility is determined by estimating an individual’s CVD risk using prediction equations, most of which have been developed using data from high-income countries^8–11^, where the population-level distribution of risk factors and disease determinants differ significantly from that of Latin American countries. Therefore, external validation in diverse populations such as Mexico is a critical step to ensure accurate CVD risk assessment.

Mexico is one of the most populous countries in Latin America, and it faces a significant CVD burden driven primarily by the high prevalence of major CVD risk factors such as diabetes, obesity, and hypertension^12–14^. Approximately 60% of the adult population has at least one CVD risk factor, and an estimated 200,000 deaths were attributed to heart disease and stroke alone in 2023^15^. Despite widespread recommendations for the use of CVD risk prediction models and their central role in guiding treatment decisions, only two such equations have included data from Mexican participants^10,16^, and none have been externally validated in a Mexican cohort for prediction of cardiovascular outcomes. Here, we aim to perform the external validation of Globorisk, Globorisk-LAC, Framingham, SCORE2, and WHO CVD risk scores for fatal CVD events, using data from adults ≥40 years enrolled in the Mexico City Prospective Study (MCPS). Additionally, we aimed to recalibrate the Globorisk fatal model to improve prediction of CVD fatal outcomes in Mexican population.

## METHODS

### Data source

We analyzed data from participants enrolled in the MCPS, a prospective, population-based cohort study with a baseline survey conducted from 1998 to 2004^17^. Full details on recruitment, procedures, and follow-up have been described previously. Briefly, households from two urban districts of Mexico City (Coyoacán and Iztapalapa) were visited, and every adult ≥35 years was invited to participate. In total, 159,517 individuals were recruited, and data regarding sociodemographic, lifestyle, and health-related information was collected by trained nurses through electronic questionnaires. Standardized measurements of height, weight, waist circumference (WC), hip circumference (HC), and sitting blood pressure (BP) were obtained using calibrated instruments. Additionally, a non-fasting 10 mL blood sample was collected from each participant for subsequent analysis. Blood samples were shipped to the University of Oxford for analysis and storage. Assays of HbA1c were performed from buffy coat samples in the Clinical Trial Service Unit and Epidemiological Studies Unit’s Wolfson laboratory. Available plasma samples from the baseline examination were analyzed using nuclear magnetic resonance (NMR) spectroscopy at Nightingale Health Ltd (Kuopio, Finland) and the Clinical Trial Service Unit’s Wolfson Laboratory, quantifying over 249 plasma biomarkers including total cholesterol, glucose, and HDL cholesterol. The study protocol was approved by the corresponding ethics committees at the Mexican Ministry of Health, the Mexican National Council for Science and Technology, and the University of Oxford. All study participants provided written informed consent.

### Predictors

Variables were defined with data collected during baseline examination and used according to the original description of each risk equation (**Supplementary Table 1**). Systolic blood pressure (SBP) was used in analyses as the mean of two or more available measurements. Diabetes was defined as self-reported medical diagnosis of diabetes, use of glucose-lowering medication, HbA1C ≥6.5, or non-fasting glucose of 11.1 mmol/L (≥200 mg/dL). Current smoking was determined according to self-reported smoking status at baseline examination. Total cholesterol, HDL-C, and glucose levels were obtained from NMR data. Individuals with extreme values of SBP (>250 mmHg or <90 mmHg), total cholesterol (>10 mmol/L), and BMI (>80 kg/m^2^ or <10 kg/m^2^) were excluded from the analyses.

### Outcomes

Only fatal CVD outcomes were evaluated, and non-fatal CVD outcomes were unavailable for analysis. Mortality follow-up of was done through electronic probabilistic linkage to Mexican death registries up to September 30th, 2022, and causes of death were classified by cohort personnel according to the International Classification of Diseases 10th Revision (ICD-10). Fatal CVD was defined as death from ischemic heart disease or sudden cardiac death (ICD10 codes I20–I25), or death from stroke (ICD10 codes I60–I69). All participants underwent administrative censoring at 10-years for evaluation of fatal outcomes.

### Statistical analyses

The estimation of fatal 10-year CVD risk was performed using the published coefficients for each risk equation: Framingham, Globorisk, Globorisk-LAC, WHO, and SCORE2 (**Supplementary Table 1**)^8–11,16,18^, including both office and laboratory based models, when available. Both fixed-time (at successive years of follow-up) and time-range (across the full follow-up period) assessment of discrimination and calibration were performed. Time-range discrimination was evaluated using Harrell’s C statistic, which quantifies concordance as the proportion of comparable participant pairs in which the individual with longer observed survival had a lower predicted risk of the outcome^19^. Fixed-point discrimination was evaluated using the time-dependent area under the receiver operating characteristic curve (AUROC)^19^. For both measures, values close to 1 indicate good discrimination ability, whereas values close to 0.5 indicate poor discrimination.

Calibration was evaluated using three complementary methods of increasing robustness, as described elsewhere^19^. First, we assessed mean calibration by calculating the ratio of the observed 10-year survival (estimated using the Kaplan-Meier estimate) to the average predicted risk, where a ratio close to 1 indicates good agreement between predicted and observed event rates. Second, we evaluated weak calibration by fitting a Cox regression model with the prognostic index as the only covariate. The resulting regression coefficient (calibration slope) should ideally be close to 1, and values below or above 1 indicate overestimation or underestimation of risk, respectively. Finally, we assessed moderate calibration by examining the agreement between predicted and observed risks across the full range of predicted probabilities. A secondary Cox model was fitted using the log(–log) transformed predicted survival probabilities as the predictor, modeled with restricted cubic splines to allow for non-linear effects. This approach allows visual inspection of the relationship between predicted and observed risks without assuming a linear calibration slope. The resulting smoothed calibration curve was then plotted against the predicted risk from the original model. Calibration is considered adequate when the curve closely follows a 45-degree line. Importantly, because we were unable to evaluate the composite outcome used in most risk equations (i.e. fatal and non-fatal CVD events), calibration estimates are likely subject to significant bias.

Finally, we recalibrated the Globorisk-fatal model separately for men and women by dividing participants into deciles of the original predicted risk within each sex. For each decile, we estimated the observed 10-year survival using the Kaplan–Meier method and then fitted a weighted least-squares linear calibration model on the complementary log–log scale, regressing the transformed observed survival on the transformed mean model survival with delta-method weights derived from the Kaplan–Meier variances. Recalibrated individual risks were obtained by applying the fitted intercept and slope to each person’s model survival on the same scale and then transforming back to risk.

## RESULTS

### Participants baseline characteristics

Of 159,517 participants recruited to MCPS, 47,255 were excluded due to missing covariate information, and 112,262 individuals with complete predictor and mortality data were included in the analysis (**Supplementary Figure 1**). Among these, 75,320 (67.09%) were women, with a median age of 52 (45-62) years, and a median age of 53 (46-63) years among men (**Table 1**). Most participants reported elementary education and residence in the Iztapalapa district (a medium-to low-income district in Mexico City). Current smoking was more prevalent among men, while diagnosed hypertension was more frequent in women, and the prevalence of diagnosed diabetes was similar between sexes. Women had slightly higher BMI, total cholesterol, and HDL-C levels compared to men. SBP and HbA1c were similar between both groups. At 10-years of follow-up there were 2,429 fatal events, including 1,667 due to myocardial infarction and 762 due to stroke. Among all events, 1,097 occurred in men and 1,332 in women. The 10-year mean CVD risk estimations varied significantly across models and between sexes. The fatal Globorisk equation showed mean risk estimates of 4.5% for both men and women. The highest median estimates were observed for the Framingham office-based equation, with 20% in men and 8.1% in women, while the lowest were estimated for the WHO stroke office-based model, with 0.82% in men and 6.53% in women (**Supplementary Table 2**).

**Table 1.** Baseline characteristics and outcomes of 112,262 participants ≥40 years from the Mexico City Prospective Study. <u>Abbreviations:</u> BMI: Body mass index, HbA1c: Glycated hemoglobin, CVD: Cardiovascular disease, MI: Myocardial infarction.

| Variable | Overall<br>N = 112,262<sup>1</sup> | Men<br>N = 36,942 | Women<br>N = 75,320 |
| --- | --- | --- | --- |
| Age (years) | 53 (45, 62) | 53 (46, 63) | 52 (45, 62) |
| Education level (%) |  |  |  |
| University | 14,714 (13%) | 7,785 (21%) | 6,929 (9.2%) |
| High school | 23,653 (21%) | 8,591 (23%) | 15,062 (20%) |
| Elementary | 57,792 (51%) | 16,900 (46%) | 40,892 (54%) |
| Other | 16,103 (14%) | 3,666 (9.9%) | 12,437 (17%) |
| Residence in Coyoacán (%) | 45,545 (41%) | 16,012 (43%) | 29,533 (39%) |
| Current smoking (%) | 33,601 (30%) | 17,648 (48%) | 15,953 (21%) |
| Diagnosed diabetes (%) | 17,390 (15%) | 5,634 (15%) | 11,756 (16%) |
| Diagnosed hypertension (%) | 26,119 (23%) | 5,931 (16%) | 20,188 (27%) |
| BMI (kg/m<sup>2</sup>) | 28.7 (25.9, 32.0) | 27.7 (25.2, 30.4) | 29.3 (26.3, 32.8) |
| Systolic BP (mmHg) | 127 (119, 138) | 128 (120, 139) | 127 (118, 138) |
| HbA1c (%) | 5.63 (5.35, 6.09) | 5.63 (5.35, 5.99) | 5.63 (5.35, 6.09) |
| Total cholesterol (mmol/l) | 4.32 (3.68, 4.97) | 4.14 (3.54, 4.77) | 4.41 (3.77, 5.06) |
| HDL cholesterol (mmol/l) | 0.99 (0.87, 1.13) | 0.92 (0.82, 1.04) | 1.03 (0.90, 1.16) |
| 10-year fatal CVD (n) | 2,429 | 1,097 | 1,332 |
| 10-year fatal MI fatal (n) | 1,667 | 792 | 875 |
| 10-year fatal stroke (n) | 762 | 305 | 457 |

<sup>1</sup>Median (Q1, Q3); n (%); n

#### Model performance – Discrimination

Time-range discrimination, assessed using Harrell’s C-statistic, was generally high across the evaluated models, although it showed sex-specific differences (**Table 2**). In men, the Globorisk fatal model presented a C-statistic of 0.800 (95%CI 0.788-0.812), while among models predicting both fatal and non-fatal outcomes, the C-statistic ranged from 0.805 (95%CI 0.783-0.827) for the WHO stroke laboratory-based model to 0.761 (0.747-0.775) for the Globorisk office-based equation. In women, model discrimination was consistently higher than in men. The Globorisk fatal model yielded a C-statistic of 0.831 (95%CI 0.821-0.841), and it ranged from 0.828 (95%CI 0.816-0.840) for the the WHO MI laboratory-based model to 0.797 (95%CI 0.785-0.809) for the Globorisk office-based equation.

**Table 2.** Discrimination of 10-year risk of fatal CVD in 112,262 participants ≥40 years of the Mexico City Prospective Study. The table shows Harrel’s c-statistics for all evaluated equations separately for men and women. <u>Abbreviations:</u> CVD: Cardiovascular disease; MI: Myocardial infarction; WHO: World Health Organization; SCORE2: Systematic Coronary Risk Evaluation 2; LAC: Latin America and the Caribbean.

| Model | C-statistic (95%CI) |  |
| --- | --- | --- |
|  | Women (n=75,320) | Men (n=36,942) |
| GloboRisk fatal | 0.831 (0.821-0.841) | 0.800 (0.788-0.812) |
| GloboRisk laboratory-based | 0.823 (0.813-0.833) | 0.792 (0.780-0.804) |
| GloboRisk office-based | 0.797 (0.785-0.809) | 0.761 (0.747-0.775) |
| GloboRisk-LAC laboratory-based | 0.824 (0.814-0.834) | 0.799 (0.787-0.811) |
| GloboRisk-LAC office-based | 0.804 (0.792-0.816) | 0.773 (0.759-0.787) |
| Framingham laboratory-based | 0.806 (0.796-0.816) | 0.777 (0.763-0.791) |
| Framingham office-based | 0.817 (0.807-0.827) | 0.779 (0.767-0.791) |
| WHO MI laboratory-based | 0.828 (0.816-0.840) | 0.793 (0.777-0.809) |
| WHO stroke laboratory-based | 0.796 (0.782-0.810) | 0.756 (0.740-0.772) |
| WHO MI office-based | 0.824 (0.806-0.842) | 0.805 (0.783-0.827) |
| WHO stroke office-based | 0.807 (0.787-0.827) | 0.791 (0.767-0.815) |
| SCORE2 | 0.821 (0.805-0.837) | 0.789 (0.771-0.807) |

Fixed-time discrimination, assessed using ROC and AUROC at successive annual time points, showed similar patterns of high performance across models (**Figure 1** and **Supplementary Table 3** and **4**). In men, AUROC values for the Globorisk fatal model ranged from 0.787 (95%CI 0.735-0.838) at year 1 to 0.815 (95%CI 0.803-0.828) at year 10. In contrast, among women, the Globorisk fatal model showed higher performance throughout follow-up, with AUROC values ranging from 0.819 (95%CI 0.808-0.829) to 0.840 (95%CI 0.837-0.842). The WHO stroke laboratory-based model and SCORE2 also presented AUROC values above 0.800 across all time points in both sexes.

**Figure 1.**
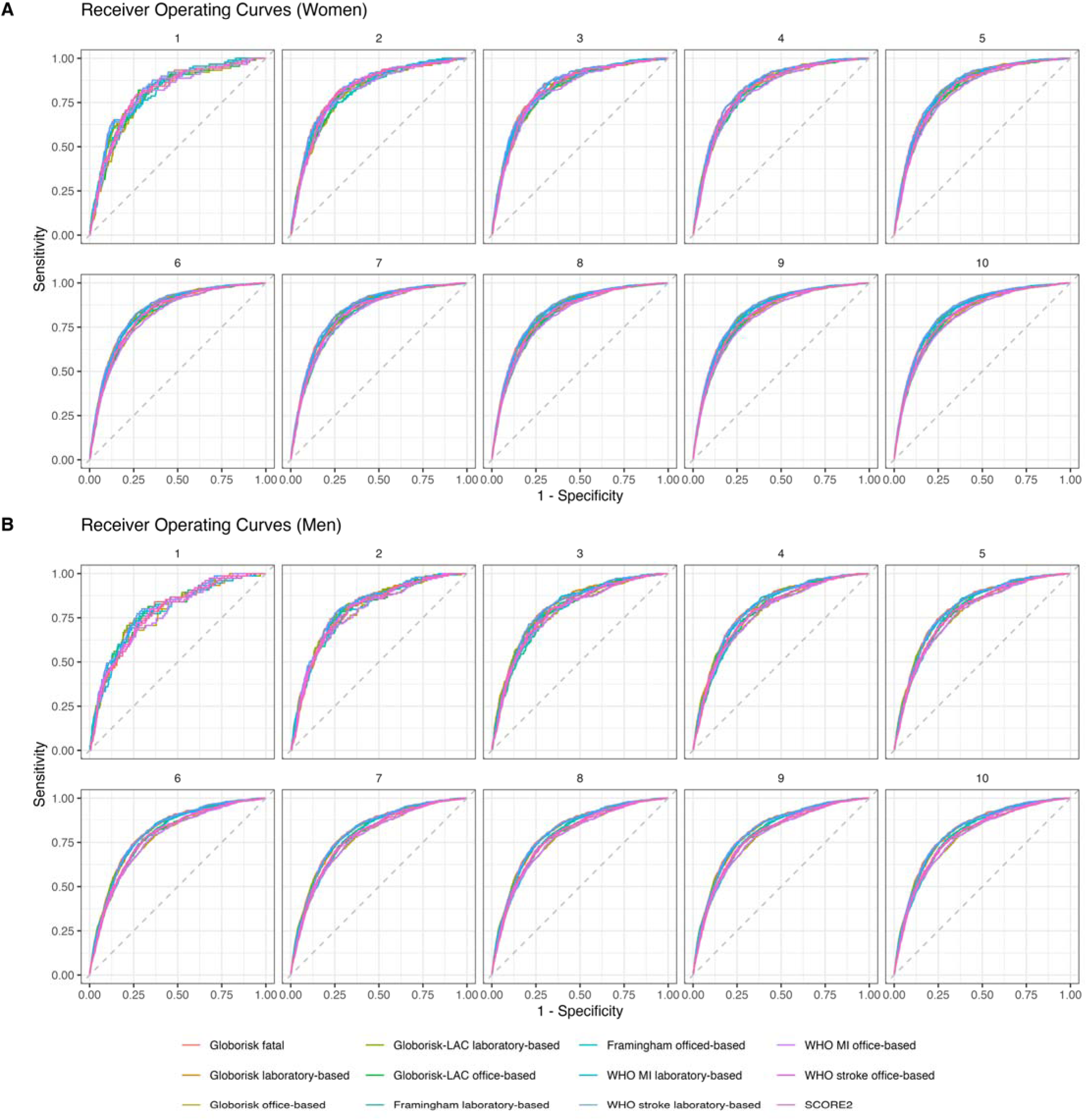
Receiver operating curves at successive annual follow-up years showing areas under the receiving operating characteristic curves (AUROCs) for prediction of 10-year risk of fatal CVD for all evaluated equations in 112,262 participants ≥40 years of the Mexico City Prospective Study, presented separately for women (A) and for men (B).

#### Model performance – Calibration

Mean calibration, which compares the average predicted risk to the observed risk providing insight into systematic over- or underestimation, also showed sex-specific differences (**Supplementary Table 5**). Despite being evaluated using the outcomes for which it was originally developed (i.e. CVD fatal events), the Globorisk fatal model significantly overestimated risk in both men and women, being more pronounced in women. All models developed for fatal- and non-fatal outcomes overestimated risk when assessed using only fatal events, resulting in systemic bias of higher predicted risks compared to the observed outcomes. Most of these models presented mean calibration values <0.50.

Weak calibration, assessed via calibration slopes, was relatively heterogeneous (**Supplementary Table 6**). In men, the Globorisk fatal model had a slope of 0.773 (95%CI 0.730-0.816), indicating overestimation at higher risks. In contrast, this model performed better in women with a slope of 0.983 (95%CI 0.937-1.029). Models predicting both fatal and non-fatal events generally exhibited slopes well above 1 in both sexes, signaling systemic overestimation at low risks and underestimation at higher predicted risks. Finally, and in line with mean and weak calibration estimates, moderate calibration showed overestimation of absolute risks with the Globorisk fatal models, particularly at higher predicted risks, in both men and women (**Figure 2**). Overestimation was most pronounced in the Framingham models, while the WHO equations, especially the MI models, showed the best agreement between predicted and observed risks (**Supplementary Figure 1**).

**Figure 2.**
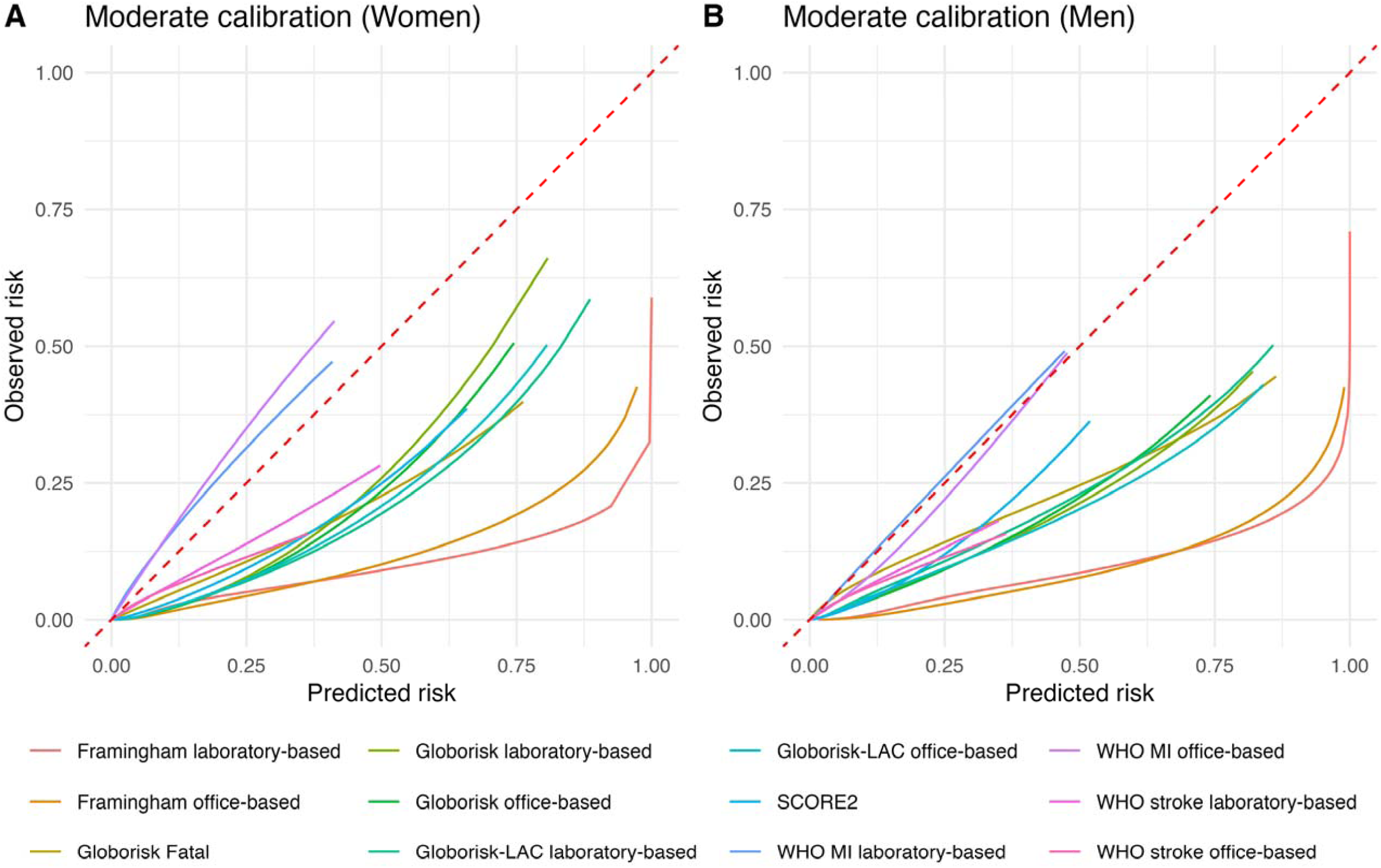
Moderate calibration plot for each evaluated equation for prediction of 10-year risk of fatal CVD in 112,262 participants ≥40 years of the Mexico City Prospective Study, presented separately for women (A) and for men (B).

#### Overall model performance

Overall performance was evaluated using the Brier score (which assesses both calibration and discrimination in a single measure of predictive accuracy, with lowest values preferred) and the scaled Brier score (which accounts for event incidence and compares the model performance to a null model, with higher percentages preferred). In women, the Globorisk fatal model demonstrated adequate overall performance with a Brier score of 0.0159 (95%CI 0.0150–0.0167) and a scaled Brier score of 9.61% (**Table 3**). The Globorisk-LAC laboratory-based and Globorisk laboratory-based models also showed strong performance with scaled scores of 8.92 and 7.96 percent, respectively. The WHO stroke models had the lowest Brier scores with 0.0060 (95%CI 0.0055–0.0066) for the laboratory-based version and 0.0061 (95%CI 0.0055–0.0066) for the office-based version, but their scaled Brier scores were among the lowest, indicating reduced relative performance given lower event rates. In men, the Globorisk fatal model showed a Brier score of 0.0274 (95%CI 0.0257–0.029) and a scaled Brier score of 6.3%, indicating good overall performance. The highest scaled score was observed for the Framingham office-based model at 9.58%, followed by the Globorisk laboratory-based model with 7.59%. In contrast, the lowest Brier scores were observed in both WHO stroke models, but with the lowest scaled scores.

**Table 3.**
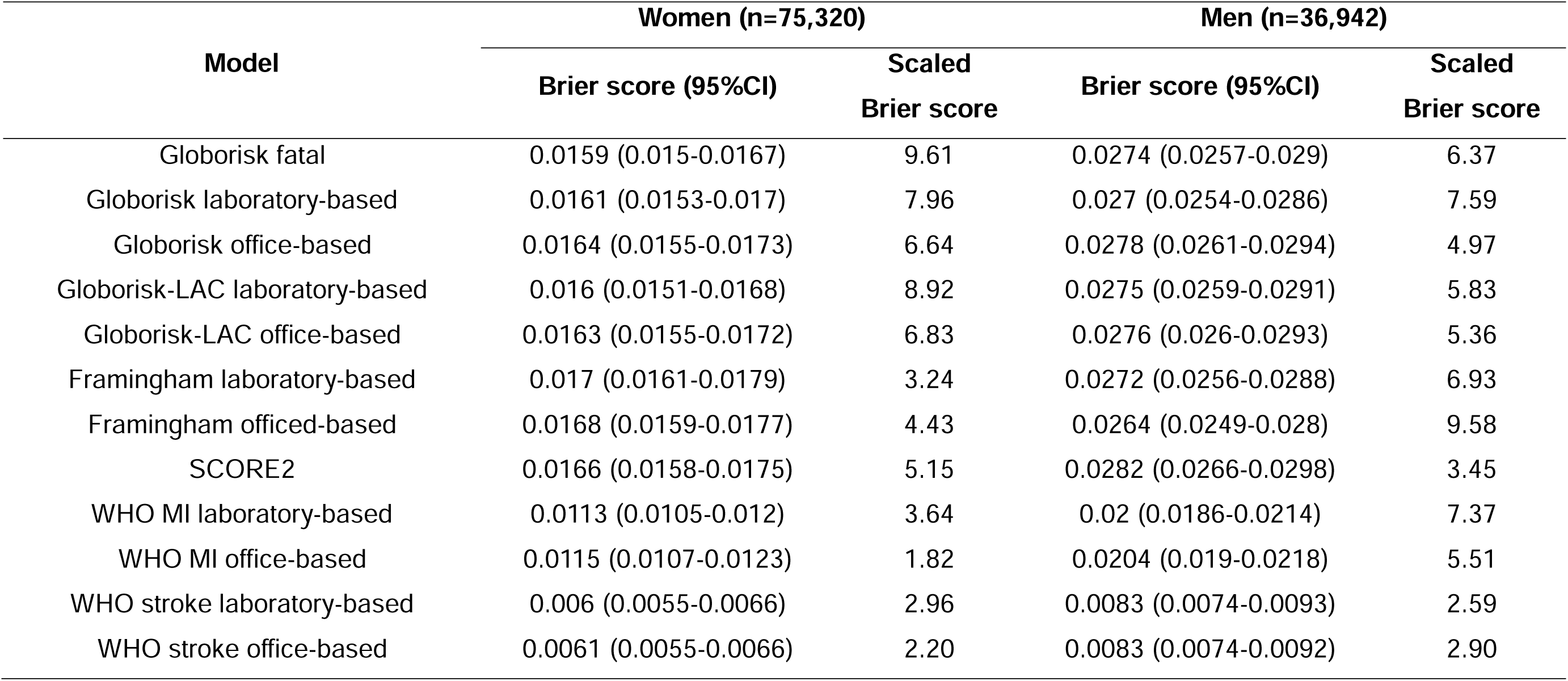
Overall model performance for prediction of 10-year risk of fatal CVD for all evaluated equations in 112,262 participants ≥40 years of the Mexico City Prospective Study. The table shows Brier scores and their respective 95% confidence intervals (95%CI), and scaled Brier scores given as percentages separately for men and women. **<u>Abbreviations:</u>** CVD: Cardiovascular disease; MI: Myocardial infarction; WHO: World Health Organization; SCORE2: Systematic Coronary Risk Evaluation 2; LAC: Latin America and the Caribbean.

#### Globorisk fatal model recalibration

The original Globorisk model substantially overestimated 10-year CVD mortality risk, particularly in women. As expected, both sexes required intercept recalibration to account for differences in baseline risk levels in the Mexican population. Notably, the recalibration slope was close to 1 in women, suggesting that relative risk gradients were well preserved, while in men the slope was below 1, indicating systematic overprediction at higher risk levels. To address this, we applied sex-specific rescaling factors (**Table 4**), recalibrating both the intercept and slope. This procedure improved risk prediction in both sexes and reduced risk overestimation (**Figure 3**).

**Figure 3.**
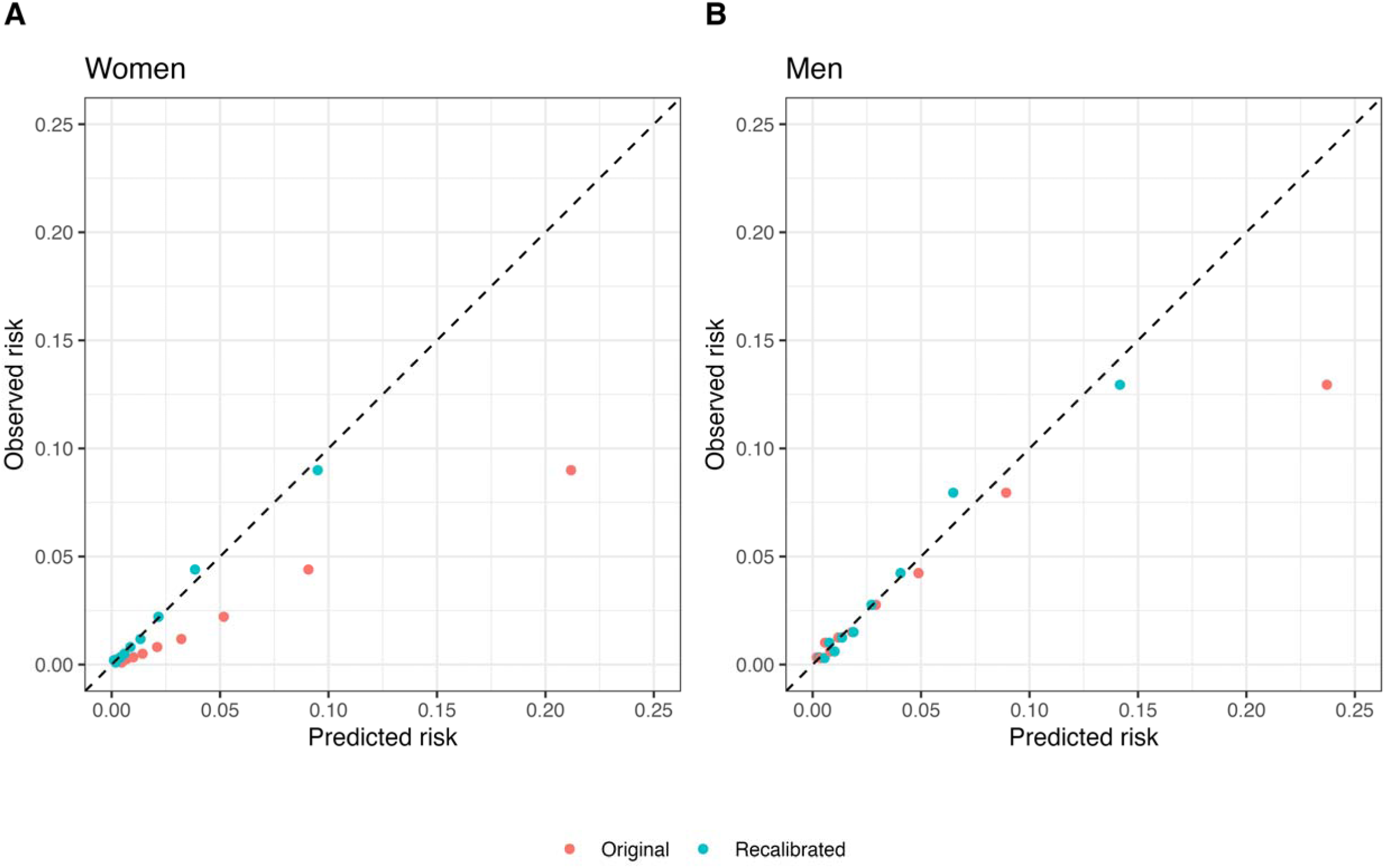
Sex-specific recalibration for 10-year CVD mortality of the Globorisk-fatal model by deciles of 10-year risk of fatal CVD in 112,262 participants ≥40 years of the Mexico City Prospective Study, presented separately for women (A) and for men (B).

**Table 4.** Sex-specific rescaling factors used for recalibration of the 10-year fatal Globorisk model to the Mexican population. To recalibrate an individual prediction: (1) compute the model survival: *S = 1 – Predicted risk*. (2) Apply the sex-specific transformation: *n = Rescaling factor 1 + Rescaling factor 2 x log(–log(S)).* (3) Transform back to survival: *S_calib_ = exp(–exp(n))* and (4) then back to risk: *r_calib_ = 1 – S_calib_*.

| Sex | Rescaling factor 1<br>(intercept, $\alpha$) | Rescaling factor 2<br>(slope, $\beta$) |
| --- | --- | --- |
| Women | -0.8820 | 1.0023 |
| Men | -0.8639 | 0.7746 |

## DISCUSSION

We performed the external validation of several widely used cardiovascular risk prediction models in a large population-based cohort from Mexico City, focusing on their ability to predict fatal CVD events over a 10-year period. In general, we found adequate discrimination across every evaluated model, though there was significant heterogeneity in calibration and overall performance, with sex-specific differences. Models originally developed for combined fatal and non-fatal outcomes performed similarly in terms of discrimination, but lack of data on non-fatal outcomes in MCPS, and the models’ subsequent assessment on fatal outcomes alone, led to systematic risk overestimation and poor calibration estimates. The Globorisk fatal model, assessed using the outcomes for which it was originally developed, showed good discrimination for both sexes and particularly strong performance in women across all metrics. However, it consistently overestimated risk, particularly at higher predicted estimates; sex-specific recalibration of this score resulted in overall predictive improvements and significant decreases in risk overestimation. These findings strengthen the notion that, even with models that have a recalibration phase during development, further adjustment to the target population is needed, taking into consideration the specific prevalence and distribution of risk factors.

Previous studies have suggested that relative risk associations for CVD risk prediction models tend to have similar magnitudes across different populations^20,21^. However, absolute risks may vary substantially due to geographic variation in risk factor levels, which typically reflect underlying epidemiological and socioeconomic patterns within a given population^22,23^. Moreover, evidence consistently demonstrates strong associations between socioeconomic status, risk factor exposure, and worsening CVD outcomes, as well as different attributable CVD mortality around the world^24–26^. Nonetheless, most CVD risk models have been developed using data from high-income countries, largely due to greater data availability. Even though recent models (i.e. SCORE2, Globorisk, Globorisk-LAC, and WHO Risk Charts) have attempted to incorporate population-level variation, risk estimation must be assessed in the target population to ensure adequate clinical decision-making^27^. This is relevant as risk estimations have a direct influence on treatment initiation and patient counseling, and in limited-resource settings, it also relates to the adequate allocation of care.

Several models have been externally validated in diverse populations, generally with heterogeneous results. The Framingham score has undergone extensive validation, usually showing risk overestimation across American, European, and Asian populations^22,28–30^. In Latin America, a recent study in Colombia found the Framingham score to overestimate risk by as much as 72%^31^, and an analysis of the Mexico City Diabetes Study also found significant overestimation, although the number of events and cohort size were limited^32^. Similarly, and in line with previous evidence, our results suggest high overestimation of predicted risks, which signal a need for recalibration, or to choose another CVD risk score.

The Globorisk model, originally developed for fatal and then for fatal plus non-fatal outcomes, has been externally validated in a few diverse populations. In a study with data from Dutch general practitioners, Globorisk significantly underestimated risk in the general population with poor calibration estimations^33^. In contrast, an analysis performed in a rural Indian cohort, found adequate discrimination and calibration, being the best model out of all the evaluated equations^34^. Interestingly, the external validation of Globorisk-LAC in a Colombian cohort found adequate discrimination but significant overprediction, despite being developed for Latin American populations^31^. Our findings appear to be similar, with adequate discrimination but significant miscalibration, which suggest the need for additional recalibration to specific Latin American countries. Finally, the SCORE2 model has been validated in European and non-European populations. SCORE2’s performance across 86 cohorts from 22 countries (European, North American, and Asian) found up to 52% of risk overestimation^22^. Similarly, in the previously mentioned Colombian cohort, SCORE2 presented adequate discrimination and moderate overestimation of risk with sex-specific differences, which is similar to the findings in our analysis^31^.

Our study has several strengths. First, it is the first study to simultaneously evaluate the predictive performance of multiple widely used CVD risk models in a large, population-based cohort from Mexico. The use of a well-characterized dataset with long-term follow-up and verified mortality outcomes adds robustness to our analysis. Moreover, the inclusion of both fatal outcome-specific models and models developed for combined fatal and non-fatal CVD outcomes allows for comparative insights into their applicability in settings with limited outcome data. Our stratified performance metrics by sex also provide important information on differential model behavior according to varying risks between men and women. However, there are also important limitations. The analysis was restricted to fatal CVD events due to the unavailability of non-fatal outcome data in MCPS, which significantly limits the generalizability of our findings for models originally developed to predict combined outcomes. Although the cohort is largely representative of urban Mexico City residents, it may not reflect the broader national population or rural settings, where risk factor distributions and healthcare access may differ. Lastly, we performed recalibration for the Globorisk fatal model, a necessary next step to enhance the predictive accuracy of existing models in the Mexican context, and which provides the first recalibrated model specific for our population of a widely applicable predictive model.

## CONCLUSION

In conclusion, our findings underscore the importance of externally validating cardiovascular risk prediction models within the populations in which they are intended to be applied, as well as the relevance of recalibration in improving predictions for CVD risk models. While every model generally demonstrated good discriminatory ability in this large urban Mexican cohort, most significantly overestimated absolute risk, highlighting the need for local recalibration or the development of region-specific tools. Additionally, we recalibrated the Globorisk fatal model for Mexican population, demonstrating improved risk estimation for fatal CVD outcomes. As risk predictions are central to clinical decision-making and resource allocation, particularly in limited-resource settings, improving their accuracy is essential for effective prevention strategies in Latin America and other low- and middle-income regions. Future studies with adequate outcomes assessment for non-fatal outcomes should focus on recalibrating existing models to broaden their clinical applicability in Mexican contexts.

## Data Availability

Data from the Mexico City Prospective Study are available to bona fide researchers. For more details, the study's Data and Sample Sharing policy may be downloaded (in English or Spanish) from https://www.ctsu.ox.ac.uk/research/mcps. Available study data can be examined in detail through the study's Data Showcase, available at https://datashare.ndph.ox.ac.uk/mexico/. Code and Supplementary Materials and Methods are available for reproducibility of results at https://github.com/oyaxbell/cvd_risk_scores_mcps.

## ETHICS APPROVAL AND CONSENT TO PARTICIPATE

The study was approved by Ethics Committees at the Mexican Ministry of Health, the Mexican National Council for Science and Technology, and the University of Oxford, UK. All participants provided written informed consent.

## CONSENT FOR PUBLICATION

Not applicable

## AUTHOR CONTRIBUTIONS

Establishing the cohort: JBC, PKM, JAD and RTC. Obtaining funding: JBC, PKM, JAD, RTC and OYBC. Data acquisition, analysis, or interpretation of data: JPE, DRG, JPDS, KBCH, LACQ, GDL, CAFM, AML, MRBA, JBC, PKM, RTC, JAD, JAS, NEAV. Drafting first version of manuscript: CAFM, OYBC. Critical revision of the report for important intellectual content: All authors. All authors have seen and approved the final version and agreed to its publication. Each author contributed important intellectual content during manuscript drafting or revision and accepts accountability for the overall work by ensuring that questions pertaining to the accuracy or integrity of any portion of the work are appropriately investigated and resolved.

## ACKNOWLEDGMENTS

This project was registered and approved by the Research Committee at Instituto Nacional de Geriatría, project number DI-PI-009/2024. JPE, CAFM, and JPDS are enrolled at the PECEM Program of the Faculty of Medicine at UNAM and are supported by SECIHTI. JAS was supported by Grant Number K23DK135798 from the NIH/NIDDK and by the Massachusetts General Hospital Executive Committee and Center for Diversity and Inclusion Physician-Scientist Development Award. The authors thank the participants for their willingness to take part in this prospective study 20 years ago. This research was conducted using Mexico City Prospective Study (MCPS) data obtained through an open-access data request (application number 2022-012).

