## Supplementary Material for "External validation and recalibration of office- and laboratory-based cardiovascular risk scores for prediction of 10-year risk of fatal cardiovascular disease in 112,262 adults in Mexico City"

### Content Table

### SUPPLEMENTARY FIGURES

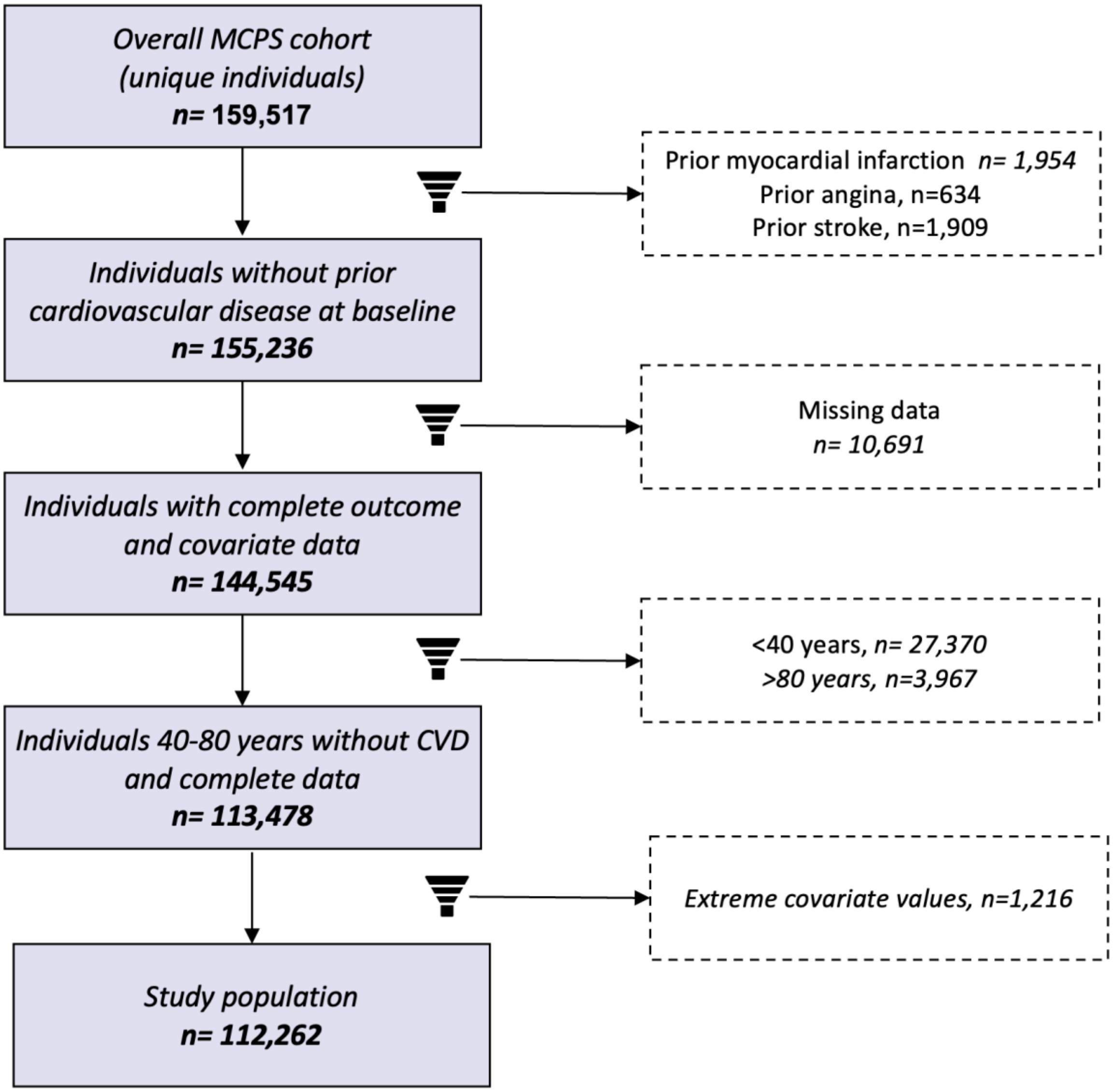

Supplementary Figure 1. Flowchart of participants from the Mexico City Prospective Study outlining reasons for exclusion from the present analysis.

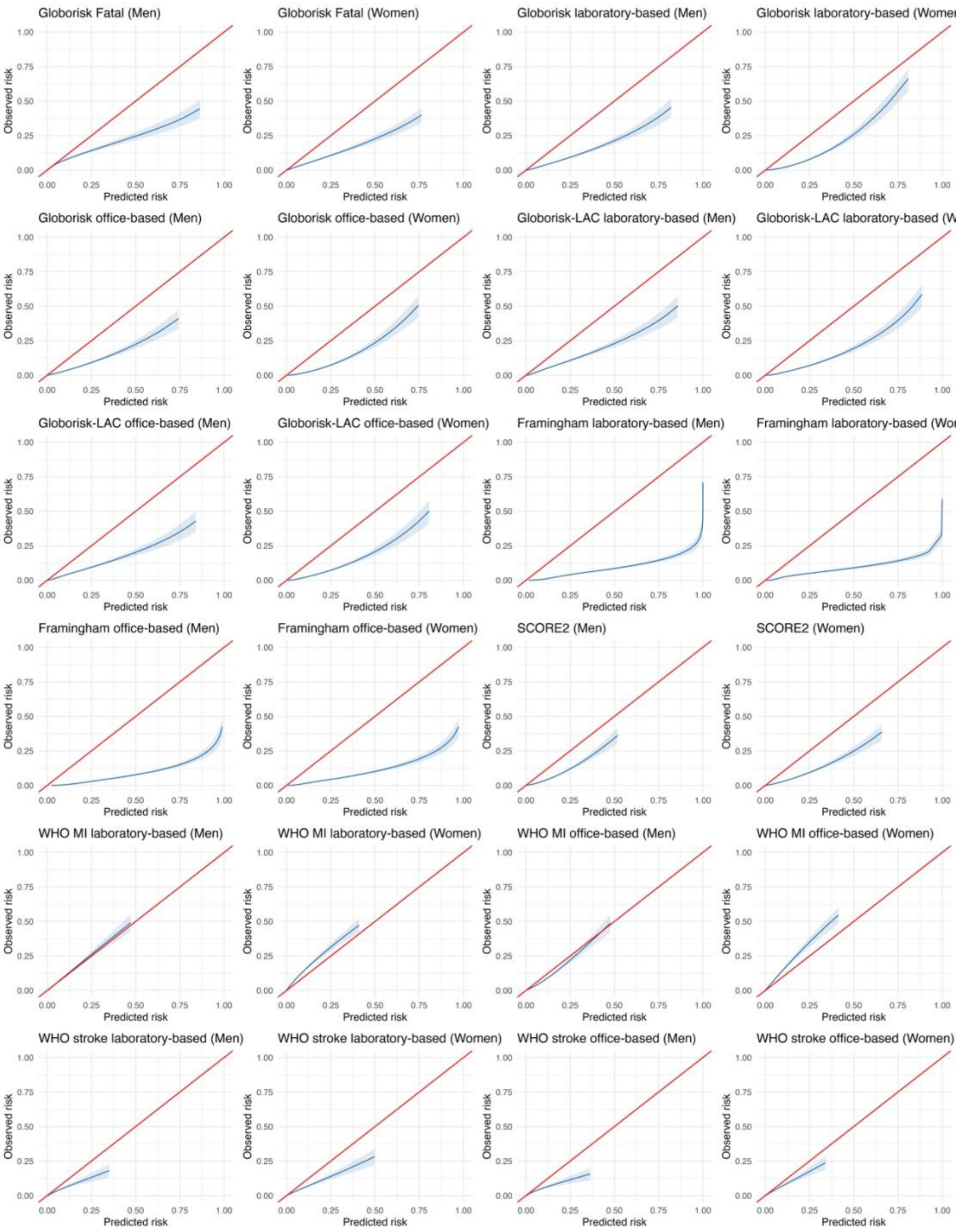

<u>*Supplementary Figure 2*</u>**.** Calibration curves  with 95% confidence intervals for all evaluated equations for risk of 10-year fatal CVD in 112,262 adults from the Mexico City Prospective Study.

**SUPPLEMENTARY TABLES**

<u>*Supplementary Table 1*</u>**.** Description of cardiovascular disease (CVD) risk prediction models included in this external validation study.

| Authors | Year | Score name | Country | Development Population | Model Outcome | Model Type | Predictors | Modelling Approach |
|---|---|---|---|---|---|---|---|---|
| D'Agostino et. al. (6) | 2008 | Framingham | USA | Framingham Heart Study and Framingham Offspring Study | Fatal/non-fatal CVD | Lab-based | Age, total cholesterol, HDL cholesterol, SBP, SBP with treatment, smoking and diabetes | Cox proportional hazards |
| | | | | | | Office-based | Age, BMI, SBP, SBP with treatment, smoking and diabetes | |
| Hajifathalian, Ueda et. al. | 2015 | Globorisk | USA | 8 prospective cohort studies | Fatal CVD | Lab-based | Age, sex, total cholesterol, SBP, smoking and diabetes | Cox proportional hazards |
| Ueda et. al. (7) | 2017 | Globorisk | USA | 8 prospective cohort studies | Fatal/non-fatal CVD | Lab-based | Age, sex, total cholesterol, SBP, smoking and diabetes | Cox proportional hazards |
| | | | | | | Office-based | Age, sex, SBP, smoking and diabetes | |
| CC-LAC (3) | 2022 | Globorisk-LAC | 6 Latin American and Caribbean countries | 9 prospective cohort studies | Fatal/non-fatal CVD | Lab-based | Age, sex, total cholesterol, SBP, smoking and diabetes | Cox proportional hazards |
| | | | | | | Office-based | Age, sex, BMI and smoking | |
| WHO CVD Risk Chart Working Group (8) | 2019 | WHO | >30 countries | Emerging Risk Factors Collabortion (85 cohorts) | Fatal/non-fatal coronary heart disease and stroke | Lab-based | Age, total cholesterol, SBP, smoking and diabetes | Cox proportional hazards |
| | | | | | | Office-based | Age, BMI, SBP and smoking | |

| Authors | Year | Score name | Country | Development Population | Model Outcome | Model Type | Predictors | Modelling Approach |
|---|---|---|---|---|---|---|---|---|
| SCORE2 working group and ESC Cardiovascular risk collaboration (9) | 2021 | SCORE2 | 13 European countries | 45 cohorts | Fatal/non-fatal CVD | Lab-based | Age, total cholesterol, HDL cholesterol, SBP, smoking and diabetes | Fine and Gray |

CC-LAC: Cohorts Consortium of Latin American and the Caribbean

*Supplementary Table 2***.** Median and interquartile range of predicted 10-year risk of CVD for all evaluated equations in 112,262 adults from the Mexico City Prospective Study.

| **Model** | **Predicted risk (%), Median (25th-75th percentile)** | |
|---|---|---|
| | **Women (n=75,320)** | **Men (n=36,942)** |
| Globorisk fatal | 1.72 (0.69–5.09) | 1.45 (0.57–4.82) |
| Globorisk laboratory-based | 6.15 (3.31–11.60) | 5.67 (2.91–12.22) |
| Globorisk office-based | 7.20 (4.28–12.09) | 6.94 (3.81–12.77) |
| Framingham laboratory-based | 6.68 (3.75–12.55) | 16.38 (9.64–28.27) |
| Framingham office-based | 8.05 (4.36–15.86) | 20.03 (11.79–34.05) |
| WHO MI laboratory-based | 0.79 (0.27–2.36) | 3.71 (1.93–7.58) |
| WHO stroke laboratory-based | 0.91 (0.35–2.74) | 1.65 (0.72–4.05) |
| WHO MI office-based | 1.08 (0.43–2.57) | 5.89 (3.49–10.21) |
| WHO stroke office-based | 0.82 (0.36–2.16) | 1.53 (0.73–3.69) |
| SCORE2 | 3.84 (1.90–8.23) | 6.53 (3.86–11.98) |
| Globorisk-LAC laboratory-based | 5.98 (3.40–11.22) | 4.99 (2.59–10.40) |
| Globorisk-LAC office-based | 6.52 (3.87–11.34) | 5.80 (3.01–11.56) |

_Supplementary Table 3_**.** Time-dependent area under the receiver operating characteristic curve for prediction of 10-year risk of CVD for all evaluated equations in 36,942 adult men from the Mexico City Prospective Study for each of the 10 years of follow-up.

| Years | Globorisk fatal | Globorisk laboratory-based | Globorisk office-based | Globorisk-LAC laboratory-based | Globorisk-LAC office-based | Framingham laboratory-based | Framingham officed-based | WHO MI laboratory-based | WHO stroke laboratory-based | WHO MI office-based | WHO stroke office-based | SCORE2 |
|---|---|---|---|---|---|---|---|---|---|---|---|---|
| 1 | 0.787 (0.735-0.838) | 0.78 (0.728-0.832) | 0.767 (0.715-0.818) | 0.792 (0.741-0.842) | 0.779 (0.728-0.83) | 0.785 (0.735-0.836) | 0.789 (0.739-0.839) | 0.778 (0.726-0.83) | 0.794 (0.745-0.844) | 0.765 (0.714-0.817) | 0.78 (0.73-0.83) | 0.775 (0.725-0.824) |
| 2 | 0.81 (0.777-0.843) | 0.801 (0.768-0.835) | 0.779 (0.743-0.815) | 0.808 (0.776-0.841) | 0.791 (0.756-0.826) | 0.792 (0.759-0.826) | 0.798 (0.764-0.831) | 0.8 (0.766-0.833) | 0.81 (0.778-0.843) | 0.778 (0.742-0.815) | 0.794 (0.759-0.829) | 0.792 (0.759-0.826) |
| 3 | 0.812 (0.786-0.837) | 0.805 (0.779-0.831) | 0.776 (0.748-0.804) | 0.809 (0.784-0.834) | 0.785 (0.759-0.812) | 0.785 (0.759-0.811) | 0.791 (0.765-0.816) | 0.803 (0.777-0.828) | 0.808 (0.783-0.832) | 0.774 (0.746-0.802) | 0.788 (0.761-0.815) | 0.786 (0.759-0.813) |
| 4 | 0.809 (0.789-0.83) | 0.802 (0.781-0.823) | 0.771 (0.748-0.793) | 0.806 (0.785-0.826) | 0.781 (0.759-0.803) | 0.784 (0.763-0.805) | 0.786 (0.766-0.807) | 0.801 (0.78-0.822) | 0.804 (0.784-0.824) | 0.769 (0.747-0.792) | 0.784 (0.762-0.806) | 0.783 (0.761-0.805) |
| 5 | 0.81 (0.792-0.828) | 0.802 (0.783-0.82) | 0.77 (0.751-0.79) | 0.807 (0.789-0.825) | 0.781 (0.762-0.801) | 0.789 (0.771-0.808) | 0.791 (0.773-0.809) | 0.802 (0.783-0.82) | 0.806 (0.788-0.824) | 0.77 (0.75-0.79) | 0.784 (0.765-0.803) | 0.782 (0.762-0.801) |
| 6 | 0.812 (0.796-0.829) | 0.804 (0.788-0.821) | 0.77 (0.752-0.788) | 0.809 (0.793-0.825) | 0.781 (0.764-0.799) | 0.791 (0.774-0.807) | 0.791 (0.774-0.807) | 0.805 (0.789-0.822) | 0.808 (0.792-0.824) | 0.769 (0.751-0.788) | 0.784 (0.766-0.801) | 0.782 (0.764-0.8) |
| 7 | 0.809 (0.794-0.825) | 0.801 (0.785-0.816) | 0.769 (0.752-0.785) | 0.807 (0.791-0.822) | 0.78 (0.764-0.797) | 0.787 (0.771-0.803) | 0.788 (0.773-0.804) | 0.802 (0.786-0.817) | 0.806 (0.79-0.821) | 0.769 (0.752-0.786) | 0.783 (0.767-0.799) | 0.779 (0.763-0.796) |
| 8 | 0.812 (0.798-0.826) | 0.803 (0.789-0.818) | 0.771 (0.756-0.787) | 0.81 (0.796-0.824) | 0.784 (0.769-0.799) | 0.789 (0.774-0.803) | 0.79 (0.775-0.804) | 0.804 (0.789-0.818) | 0.809 (0.795-0.823) | 0.771 (0.755-0.787) | 0.786 (0.771-0.801) | 0.782 (0.766-0.797) |
| 9 | 0.815 (0.802-0.828) | 0.806 (0.793-0.82) | 0.774 (0.759-0.789) | 0.813 (0.799-0.826) | 0.786 (0.772-0.8) | 0.791 (0.777-0.804) | 0.792 (0.779-0.806) | 0.807 (0.794-0.821) | 0.811 (0.798-0.824) | 0.774 (0.759-0.789) | 0.789 (0.775-0.803) | 0.784 (0.77-0.799) |
| 10 | 0.815 (0.803-0.828) | 0.806 (0.794-0.819) | 0.774 (0.76-0.788) | 0.813 (0.801-0.826) | 0.787 (0.774-0.8) | 0.791 (0.778-0.803) | 0.792 (0.78-0.805) | 0.807 (0.795-0.82) | 0.812 (0.8-0.825) | 0.774 (0.761-0.788) | 0.789 (0.776-0.803) | 0.784 (0.77-0.798) |

Supplementary Table 4. Time-dependent area under the receiver operating characteristic curve for prediction of 10-year risk of CVD for all evaluated equations in 75,320 adult women from the Mexico City Prospective Study for each of the 10 years of follow-up.

| Years | Globorisk fatal | Globorisk laboratory-based | Globorisk office-based | Globorisk-LAC laboratory-based | Globorisk-LAC office-based | Framingham laboratory-based | Framingham officed-based | WHO MI laboratory-based | WHO stroke laboratory-based | WHO MI office-based | WHO stroke office-based | SCORE2 |
|---|---|---|---|---|---|---|---|---|---|---|---|---|
| 1 | 0.819 (0.808-0.829) | 0.81 (0.799-0.82) | 0.784 (0.773-0.795) | 0.812 (0.802-0.823) | 0.795 (0.784-0.806) | 0.806 (0.796-0.816) | 0.798 (0.787-0.809) | 0.816 (0.806-0.826) | 0.82 (0.81-0.831) | 0.79 (0.779-0.801) | 0.806 (0.795-0.817) | 0.808 (0.797-0.818) |
| 2 | 0.832 (0.825-0.839) | 0.822 (0.815-0.829) | 0.8 (0.793-0.807) | 0.823 (0.816-0.829) | 0.81 (0.803-0.817) | 0.806 (0.798-0.813) | 0.811 (0.804-0.817) | 0.824 (0.818-0.831) | 0.833 (0.826-0.839) | 0.802 (0.795-0.809) | 0.819 (0.812-0.826) | 0.82 (0.813-0.827) |
| 3 | 0.839 (0.834-0.845) | 0.83 (0.824-0.835) | 0.808 (0.802-0.814) | 0.832 (0.827-0.837) | 0.816 (0.81-0.822) | 0.816 (0.81-0.821) | 0.825 (0.82-0.831) | 0.833 (0.827-0.838) | 0.841 (0.836-0.846) | 0.807 (0.802-0.813) | 0.823 (0.817-0.828) | 0.823 (0.818-0.829) |
| 4 | 0.839 (0.835-0.844) | 0.83 (0.826-0.835) | 0.81 (0.806-0.815) | 0.832 (0.827-0.836) | 0.818 (0.813-0.822) | 0.815 (0.81-0.819) | 0.826 (0.822-0.83) | 0.831 (0.827-0.835) | 0.84 (0.836-0.844) | 0.807 (0.803-0.812) | 0.822 (0.818-0.827) | 0.822 (0.817-0.826) |
| 5 | 0.842 (0.838-0.845) | 0.833 (0.829-0.837) | 0.809 (0.805-0.813) | 0.835 (0.832-0.839) | 0.817 (0.813-0.821) | 0.819 (0.815-0.823) | 0.829 (0.825-0.833) | 0.833 (0.829-0.837) | 0.842 (0.838-0.845) | 0.806 (0.802-0.81) | 0.822 (0.818-0.826) | 0.821 (0.817-0.825) |
| 6 | 0.844 (0.841-0.847) | 0.837 (0.833-0.84) | 0.811 (0.807-0.815) | 0.839 (0.835-0.842) | 0.82 (0.816-0.823) | 0.822 (0.818-0.826) | 0.832 (0.828-0.835) | 0.835 (0.832-0.839) | 0.844 (0.841-0.847) | 0.807 (0.804-0.811) | 0.824 (0.82-0.827) | 0.822 (0.819-0.826) |
| 7 | 0.843 (0.84-0.846) | 0.835 (0.832-0.838) | 0.811 (0.808-0.815) | 0.836 (0.833-0.839) | 0.818 (0.815-0.821) | 0.816 (0.813-0.819) | 0.827 (0.824-0.83) | 0.835 (0.832-0.838) | 0.843 (0.84-0.846) | 0.809 (0.805-0.812) | 0.823 (0.82-0.827) | 0.822 (0.819-0.825) |
| 8 | 0.841 (0.838-0.844) | 0.833 (0.83-0.836) | 0.809 (0.806-0.812) | 0.834 (0.832-0.837) | 0.816 (0.813-0.819) | 0.815 (0.812-0.818) | 0.826 (0.823-0.829) | 0.833 (0.83-0.836) | 0.841 (0.839-0.844) | 0.806 (0.803-0.81) | 0.821 (0.818-0.824) | 0.819 (0.816-0.822) |
| 9 | 0.84 (0.838-0.843) | 0.832 (0.83-0.835) | 0.806 (0.803-0.809) | 0.833 (0.831-0.836) | 0.814 (0.811-0.817) | 0.814 (0.811-0.817) | 0.825 (0.823-0.828) | 0.832 (0.829-0.835) | 0.84 (0.838-0.843) | 0.804 (0.801-0.806) | 0.819 (0.816-0.822) | 0.817 (0.814-0.82) |
| 10 | 0.84 (0.837-0.842) | 0.832 (0.83-0.835) | 0.805 (0.802-0.808) | 0.833 (0.83-0.835) | 0.812 (0.809-0.815) | 0.814 (0.811-0.817) | 0.825 (0.823-0.828) | 0.832 (0.83-0.835) | 0.84 (0.837-0.842) | 0.802 (0.8-0.805) | 0.818 (0.815-0.82) | 0.815 (0.813-0.818) |

Supplementary Table 5. Mean and observed risk of 10-year fatal CVD, and mean calibration estimates with 95% confidence intervals for all evaluated equations in 112,262 adults from the Mexico City Prospective Study.

| Model | Women (n=75,320) | | | Men (n=36,942) | | |
|---|---|---|---|---|---|---|
| | **Mean observed risk** | **Mean predicted risk** | **Mean calibration (95% CI)** | **Mean observed risk** | **Mean predicted risk** | **Mean calibration (95% CI)** |
| Globorisk fatal | 0.018 | 0.045 | 0.408 (0.387-0.431) | 0.031 | 0.045 | 0.686 (0.646-0.727) |
| Globorisk laboratory-based | 0.018 | 0.086 | 0.211 (0.2-0.222) | 0.031 | 0.094 | 0.332 (0.313-0.352) |
| Globorisk office-based | 0.018 | 0.092 | 0.198 (0.188-0.209) | 0.031 | 0.097 | 0.321 (0.302-0.34) |
| Framingham laboratory-based | 0.018 | 0.099 | 0.184 (0.175-0.194) | 0.031 | 0.212 | 0.147 (0.138-0.156) |
| Framingham office-based | 0.018 | 0.123 | 0.148 (0.141-0.157) | 0.031 | 0.250 | 0.124 (0.117-0.132) |
| SCORE2 | 0.018 | 0.064 | 0.286 (0.271-0.302) | 0.031 | 0.089 | 0.349 (0.329-0.37) |
| Globorisk-LAC laboratory-based | 0.018 | 0.088 | 0.208 (0.197-0.219) | 0.031 | 0.083 | 0.375 (0.354-0.398) |
| Globorisk-LAC office-based | 0.018 | 0.088 | 0.207 (0.197-0.219) | 0.031 | 0.090 | 0.345 (0.325-0.366) |
| WHO MI laboratory-based | 0.012 | 0.020 | 0.602 (0.563-0.643) | 0.023 | 0.056 | 0.406 (0.379-0.435) |
| WHO stroke laboratory-based | 0.012 | 0.021 | 0.578 (0.541-0.618) | 0.023 | 0.076 | 0.297 (0.277-0.319) |
| WHO MI office-based | 0.006 | 0.022 | 0.279 (0.255-0.306) | 0.009 | 0.030 | 0.291 (0.26-0.326) |
| WHO stroke office-based | 0.006 | 0.018 | 0.349 (0.318-0.383) | 0.009 | 0.029 | 0.303 (0.271-0.339) |

*Supplementary Table 6*. Weak calibration estimates with 95% confidence intervals for all evaluated equations for risk of 10-year fatal CVD in 112,262 adults from the Mexico City Prospective Study.

| **Model** | **Men (n=36,942)** | **Women (n=75,320)** |
|---|---|---|
| | **Coefficient (95%CI)** | **Coefficient (95%CI)** |
| Globorisk fatal | 0.7732 (0.7303-0.8162) | 0.9825 (0.9365-1.0285) |
| Globorisk laboratory-based | 1.0876 (1.0247-1.1505) | 1.5137 (1.4412-1.5861) |
| Globorisk office-based | 1.1210 (1.0479-1.194) | 1.5184 (1.4411-1.5956) |
| Globorisk-LAC laboratory-based | 1.0993 (1.0389-1.1597) | 1.3950 (1.3314-1.4586) |
| Globorisk-LAC office-based | 1.0534 (0.9889-1.1178) | 1.4374 (1.3664-1.5084) |
| Framingham laboratory-based | 1.0242 (0.9683-1.08) | 0.9073 (0.8697-0.9448) |
| Framingham officed-based | 1.2353 (1.1616-1.3091) | 1.2173 (1.1597-1.2748) |
| WHO MI laboratory-based | 1.2833 (1.1933-1.3733) | 0.9941 (0.934-1.0543) |
| WHO stroke laboratory-based | 1.1417 (1.0152-1.2682) | 1.0302 (0.943-1.1174) |
| WHO MI office-based | 1.3206 (1.2153-1.4259) | 1.0092 (0.943-1.0754) |
| WHO stroke office-based | 1.0464 (0.9277-1.1651) | 1.0213 (0.9323-1.1102) |
| SCORE2 | 1.4432 (1.3354-1.5511) | 1.3072 (1.2262-1.3882) |
